# Full vaccination against COVID-19 suppresses SARS-CoV-2 delta variant and spike gene mutation frequencies and generates purifying selection pressure

**DOI:** 10.1101/2021.08.08.21261768

**Authors:** Ting-Yu Yeh, Gregory P. Contreras

## Abstract

COVID-19 vaccination resistance has become a major challenge to prevent global SARS-CoV-2 transmission. Here we report that the vaccination coverage rate is inversely correlated to the mutation frequency of the full genome (*R*^2^=0.878) and spike gene (*R*^2^=0.829) of SARS-CoV-2 delta variants in 16 countries, suggesting that full vaccination against COVID-19, with other mitigation strategies, is critical to suppress emergent mutations. Neutrality analysis of *DH* and Zeng’s *E* tests suggested that directional selection was the major driving force of delta variant evolution. To eliminate the homogenous effects (population expansion, selective sweep etc.), the synonymous (*D*_syn_) and nonsynonymous (*D*_nonsyn_) polymorphisms of the delta variant spike gene were estimated with Tajima’s *D* statistic. Both *D* ratio (*D*_nonsyn_/*D*_syn_) and Δ*D* (*D*_syn_-*D*_nonsyn_) have positive correlation with the full vaccination rate (*R*^2^= 0.723 and 0.505, respectively) in 19 countries, indicating that purifying selection pressure of SARS-CoV-2 spike gene increased as the vaccination coverage rate increased. Taken together, our data suggests that vaccination plays an important role in the purifying selection force of spike protein of SARS-CoV-2 delta variants.

## Introduction

Legally required vaccination against various infectious diseases is essential to public health policy in many countries. However, resistance to voluntary COVID-19 vaccination has emerged worldwide, including in the United States. In mid-June 2021, 33% of Americans said they were not willing to be vaccinated (Kirzinger et al, 2021). Public distrust has undermined COVID-19 vaccine acceptance, in association with a belief that the vaccine is ineffective, dangerous or compromises individual freedom (Schmelz et al, 2021). Therefore, overcoming COVID-19 vaccination resistance has become a major challenge to prevent global SARS-CoV-2 transmission.

Mutations drive genome variability, generating many different SARS-CoV-2 variants as the virus evolves to escape vaccine-mediated immunity and thereby, develop drug or vaccine resistance (Roy et al., 2020). The delta (lineage B1.617.2) variants were first documented in October 2020 in India and were designated variants of concern by the World Health Organization in May of 2021 due to its increased transmissibility and virulence (https://www.who.int/en/activities/tracking-SARS-CoV-2-variants/). All SARS-CoV-2 mutations result from two major mechanisms: (1) spontaneous substitution/deletion of nucleotides, and (2) RNA recombination (Roy et al, 2020, Yeh and Contreras, 2020, Ignatieva et al, 2021, Yeh and Contreras, 2021). Until now, it remained unclear exactly how human vaccinations affected virus selective pressure.

### Is there correlation between mutation frequency and vaccination?

To explore this question, we first analyzed the correlation between the rates of full vaccination and the point mutation frequency (*Mf*) of COVID-19 delta variants’ genome in 20 countries. Complete SARS-CoV-2 genome sequences with high coverage from June 20 to July 3 2021 in 20 countries: Australia (*N*=121), France (*N*=788), Germany (*N*=955), Indonesia (*N*=97), India (*N*=171), Ireland (*N*=617), Israel (*N*=333), Italy (*N*=642), Japan (*N*=105), Mexico (*N*=368), Netherland (*N*=456), Norway (*N*=142), Portugal (*N*=782), Singapore (*N*=131), Spain (*N*=689), Switzerland (*N*=131), Sweden (*N*=786), Turkey (*N*=428), United States (*N*=537), and United Kingdom (UK, *N*=3534) were collected from the Global Initiative on Sharing All Influenza Data (GISAID) (https://www.gisaid.org/). *Mf* was calculated as Pi / (Ln × Ns). Pi is the number of instances of polymorphism detected within the genome/locus. Ln is the nucleotide length of the genome/locus. Ns is the number of sequenced entities present in the dataset (Roy et al., 2020). The information of fully vaccinated rate on 3 July 2021 in individual counties were obtained in Our World in Data (https://ourworldindata.org/covid-vaccinations).

We found that *Mf* was logarithmically reduced as the full vaccination rate increased in 16 of 20 countries (*R*^2^=0.878, *P*<10^−7^, Figure 1A). To our knowledge, this is the first evidence suggesting that vaccinations could successfully suppress viral mutations. Since the spike protein is the target of the vaccination program, we further examined mutations of delta variant spike gene. Likewise, we found that the *Mf* of the spike gene was also logarithmically reduced as the full vaccination rate increased this time in 17 of the 20 countries (*R*^2^=0.829, *P*<10^−6^, Figure 1B). With a 10.8% vaccinated rate, *Mf* was exceptionally low in Australia, likely as a result of the strict state lockdown restrictions. In contrast, the *Mfs* are higher in Japan, Switzerland, and United States, suggesting that their social mitigation strategies have been less successful. This also suggests that he mutation frequency may be not only affected by vaccination only, but may also reflect overall control of the virus.

**Figure 1.**
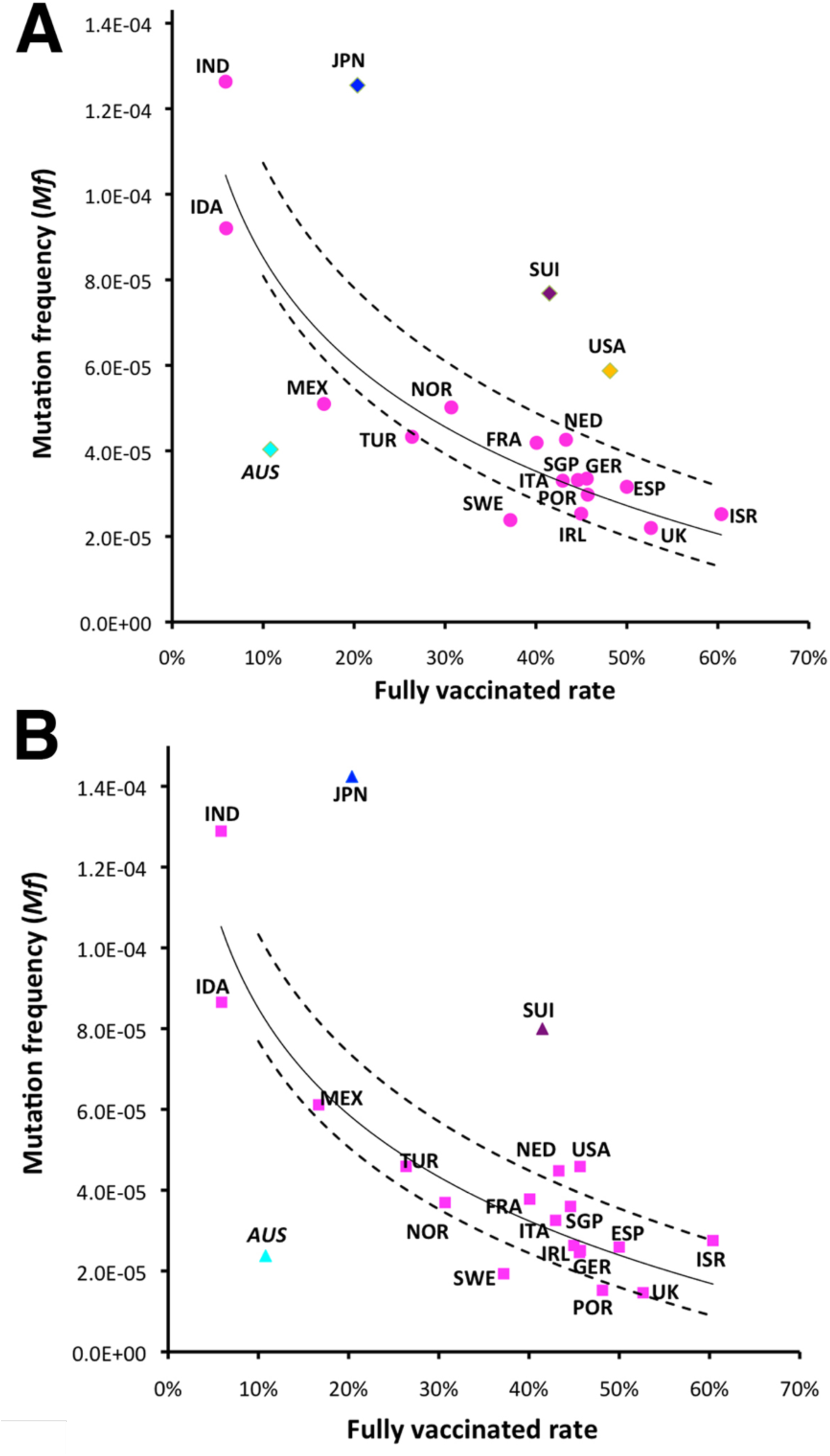
Correlation between full vaccinated rate (https://ourworldindata.org/covid-vaccinations) and mutation frequency (*Mf*) of the full genome (A) or the spike gene (B) of SARS-CoV-2 delta variants from June 20 to July 3 2021 in 20 countries: Australia (AUS), France (FRA), Germany (GER), Indonesia (IDA), India (IND), Ireland (IRL), Israel (ISR), Italy (ITA), Japan (JPN), Mexico (MEX), Netherland (NED), Norway (NOR), Portugal (POR), Singapore (SGP), Spain (ESP), Switzerland (SUI), Sweden (SWE), Turkey (TUR), United States (USA), and UK. Logarithmic regression (solid) line was draw based on 16 countries (pink) with a calculated 95% confidence interval (dashed lines). Japan, Switzerland, USA, and Australia are labeled in different colors as outliers.

### How does vaccination affect selection pressure?

SARS-CoV-2 is subject to many powerful selection pressures which may contribute to vaccine escape. Like other viruses, the delta variant appears to have been selected for its increased transmissibility regardless of vaccination. Previously we reported on the mutations and selection pressure of SARS-CoV-2 among passengers in close-quarters quarantine on the Diamond Princess cruise ship between January and March 2020 (Yeh and Contreras, 2021). Until then, it had been unclear whether COVID-19 variants were evolving randomly or under selection pressures generated by vaccination and/or other mitigation efforts.

To understand how selection pressure operates within COVID-19 virus populations, we analyzed the polymorphism of the full SARS-CoV-2 genome using neutrality tests based on the site-frequency spectrum: (1) Tajima’s *D* test, (2) normalized Wu and Fay’s *DH* test, and (3) Zeng’s *E* test, using DNASP6 software with the bat coronavirus RaTG13 as the outgroup sequence (Rozas, et al., 2017). The Tajima *D* test compares pairwise nucleotide diversity (π, the average number of nucleotide differences per site between two sequences) and total polymorphism to infer selection and/or demographic events (Tajima, 1989). *DH* test is sensitive to the changes in high-frequency variants and primarily affected by directional selection and no other driving forces, such as demographic expansion (Zeng et al., 2006). Zeng’s *E* test contrasts high and low frequency regions of frequency spectrum, so this test can detect population growth after a sweep (Zeng et al., 2006). Statistical significance of observed values of all tests was obtained by coalescent simulation with free recombination after 10,000 replicates. The probability for each test statistic was calculated as the frequency of replicates with a value lower than the observed statistic (two-tailed test) by DNASP6 (Rozas, et al., 2017).

We observed that Tajima’s *D* values were negative and significantly deviated from zero (−2.199 to -2.878, *P* < 0.05) among 19 countries, indicating an excess of variants of low frequency (Table 1). Because the negative Tajima’s *D* values could result from demographic expansion and/or purifying/positive selection, we further employed normalized *DH* (insensitive to population growth) and Zeng’s *E* test (Zeng et al., 2006). Because fixation causes quick loss of high-frequency alleles by drift, and excess of low-frequency mutations are generated, Zeng’s *E* test is very sensitive to population growth. Genomic polymorphism of these viral populations revealed significantly negative *DH* values (−1.177 to -2.791, *P*<0.05), but *E* values were not significantly different from zero (−0.04 to 0.482, *P*>0.05) (Table 1). Similar results were obtained after confining our analysis to the spike gene of SARS-CoV-2 delta variants using Tajima’ *D, DH*, and Zeng’s *E* tests (Table 1). Taken together, these data suggest that directional selection was operating in these countries, and demographic expansion was less likely the major driving force.

**Table 1.** Neutrality analysis of SARS-CoV-2 delta variants of the full genome and the viral spike gene from June 20 to July 3, 2021 in 20 countries. The statistical significance was estimated using 10,000 coalescent simulations in DNASP6. Not significant values (*P*>0.05) are indicated in italic.

| Country | Tajima's<br>*D* | *DH* | Zeng's<br>*E* | *DH* | Zeng's<br>*E* | Tajima's<br>*D* | $\Delta D$<br>($D_{\text{Nonsyn}}$<br>-$D_{\text{Syn}}$) | *D* ratio<br>($D_{\text{Nonsyn}}$<br>/$D_{\text{Syn}}$) | Ka<br>/Ks | Neutrality<br>index |
| --- | --- | --- | --- | --- | --- | --- | --- | --- | --- | --- |
| Israel | -2.401 | -2.539 | *-0.040* | -4.468 | *1.792* | -2.065 | 0.611 | 1.424 | 0.055 | 13.209 |
| France | -2.518 | -2.482 | *-0.278* | -2.973 | *0.176* | -2.482 | 0.307 | 1.144 | 0.055 | 15.612 |
| Germany | -2.800 | -2.703 | *-0.370* | -3.831 | *0.655* | -2.661 | 0.065 | 1.026 | 0.034 | 10.938 |
| Portugal | -2.645 | -2.269 | *-0.558* | -3.16 | *0.402* | -2.395 | 0.593 | 1.314 | 0.058 | 8.393 |
| Spain | -2.660 | -2.61 | *-0.283* | *-1.606* | *-0.943* | -2.530 | 0.306 | 1.143 | 0.065 | 12.986 |
| Australia | -2.578 | -1.914 | *-0.626* | *-1.949* | *-0.078* | -2.064 | 0.091 | 1.056 | 0.052 | 10.609 |
| USA | -2.657 | -2.207 | *-0.594* | -4.551 | *1.306* | -2.580 | 0.290 | 1.124 | 0.015 | 14.467 |
| Ireland | -2.833 | -2.069 | *-0.868* | -2.337 | *-0.464* | -2.659 | 0.289 | 1.126 | 0.055 | 12.385 |
| Italy | -2.533 | -2.341 | *-0.388* | -3.691 | *0.73* | -2.51 | 0.095 | 1.042 | 0.045 | 17.699 |
| Turkey | -2.451 | -2.638 | *-0.252* | -3.748 | *0.916* | -2.398 | -0.09 | 0.961 | 0.044 | 16.292 |
| Indonesia | -2.339 | -2.116 | *-0.211* | *-1.35* | *-0.342* | -1.773 | -0.381 | 0.796 | 0.055 | 11.236 |
| Japan | -2.523 | -1.801 | *-0.675* | -3.697 | *1.095* | -2.459 | -0.582 | 0.776 | 0.052 | 5.068 |
| Mexico | -2.522 | -2.532 | *-0.166* | -3.111 | *0.444* | -2.405 | -0.307 | 0.869 | 0.073 | 11.340 |
| Norway | -1.566 | -1.177 | *-0.370* | *-0.682* | *-0.732* | -1.562 | 0.359 | 1.307 | 0.054 | 9.059 |
| Switzerland | -2.560 | -2.319 | *-0.279* | -4.591 | *1.756* | -2.494 | 0.333 | 1.158 | 0.045 | 10.81 |
| Sweden | -2.473 | -2.295 | *-0.383* | -4.275 | *1.101* | -2.565 | 0.182 | 1.08 | 0.021 | 11.19 |
| Netherlands | -2.372 | -2.327 | *-0.231* | -3.811 | *0.893* | -2.48 | -0.107 | 0.956 | 0.049 | 10.64 |
| Singapore | -2.199 | -2.791 | *0.482* | -2.538 | *0.507* | -1.909 | 0.500 | 1.364 | 0.053 | 9.059 |
| India | -2.878 | -2.470 | *-0.469* | -3.961 | *0.884* | -2.826 | -0.073 | 0.793 | 0.051 | 7.102 |
| UK | -2.748 | -2.546 | *-0.540* | -3.299 | *0.168* | -2.663 | 0.355 | 1.162 | 0.013 | 32.537 |

Table 1. Neutrality analysis of SARS-CoV-2 delta variants of the full genome and the viral spike gene from June 20 to July 3, 2021 in 20 countries. The statistic significance was estimated using 10,000 coalescent simulations in DNASP6. Not significant values ( $P>0.05$ ) are indicated in italic.
|  | Full genome |  |  | Spike gene |  |  |  |  |  |  |
| --- | --- | --- | --- | --- | --- | --- | --- | --- | --- | --- |
| Country | Tajima's *D* | *DH* | Zeng's *E* | *DH* | Zeng's *E* | Tajima's *D* | $\Delta D$<br>($D_{\text{Nonsyn}}$<br>-$D_{\text{syn}}$) | *D* ratio<br>($D_{\text{Nonsyn}}$<br>/$D_{\text{syn}}$) | Ka<br>/Ks | Neutrality<br>index |
| Israel | -2.401 | -2.539 | -0.040 | -4.468 | 1.792 | -2.065 | 0.611 | 1.424 | 0.055 | 13.209 |
| France | -2.518 | -2.482 | -0.278 | -2.973 | 0.176 | -2.482 | 0.307 | 1.144 | 0.055 | 15.612 |
| Germany | -2.800 | -2.703 | -0.370 | -3.831 | 0.655 | -2.661 | 0.065 | 1.026 | 0.034 | 10.938 |
| Portugal | -2.645 | -2.269 | -0.558 | -3.16 | 0.402 | -2.395 | 0.593 | 1.314 | 0.058 | 8.393 |
| Spain | -2.660 | -2.61 | -0.283 | -1.606 | -0.943 | -2.530 | 0.306 | 1.143 | 0.065 | 12.986 |
| Australia | -2.578 | -1.914 | -0.626 | -1.949 | -0.078 | -2.064 | 0.091 | 1.056 | 0.052 | 10.609 |
| USA | -2.657 | -2.207 | -0.594 | -4.551 | 1.306 | -2.580 | 0.290 | 1.124 | 0.015 | 14.467 |
| Ireland | -2.833 | -2.069 | -0.868 | -2.337 | -0.464 | -2.659 | 0.289 | 1.126 | 0.055 | 12.385 |
| Italy | -2.533 | -2.341 | -0.388 | -3.691 | 0.73 | -2.51 | 0.095 | 1.042 | 0.045 | 17.699 |
| Turkey | -2.451 | -2.638 | -0.252 | -3.748 | 0.916 | -2.398 | -0.09 | 0.961 | 0.044 | 16.292 |
| Indonesia | -2.339 | -2.116 | -0.211 | -1.35 | -0.342 | -1.773 | -0.381 | 0.796 | 0.055 | 11.236 |
| Japan | -2.523 | -1.801 | -0.675 | -3.697 | 1.095 | -2.459 | -0.582 | 0.776 | 0.052 | 5.068 |
| Mexico | -2.522 | -2.532 | -0.166 | -3.111 | 0.444 | -2.405 | -0.307 | 0.869 | 0.073 | 11.340 |
| Norway | -1.566 | -1.177 | -0.370 | -0.682 | -0.732 | -1.562 | 0.359 | 1.307 | 0.054 | 9.059 |
| Switzerland | -2.560 | -2.319 | -0.279 | -4.591 | 1.756 | -2.494 | 0.333 | 1.158 | 0.045 | 10.81 |
| Sweden | -2.473 | -2.295 | -0.383 | -4.275 | 1.101 | -2.565 | 0.182 | 1.08 | 0.021 | 11.19 |
| Netherlands | -2.372 | -2.327 | -0.231 | -3.811 | 0.893 | -2.48 | -0.107 | 0.956 | 0.049 | 10.64 |
| Singapore | -2.199 | -2.791 | 0.482 | -2.538 | 0.507 | -1.909 | 0.500 | 1.364 | 0.053 | 9.059 |
| India | -2.878 | -2.470 | -0.469 | -3.961 | 0.884 | -2.826 | -0.073 | 0.793 | 0.051 | 7.102 |
| UK | -2.748 | -2.546 | -0.540 | -3.299 | 0.168 | -2.663 | 0.355 | 1.162 | 0.013 | 32.537 |

Selection pressure has been observed in unvaccinated SARS-CoV-2 populations, such as in the aforementioned Diamond Princess shipboard lockdown (Yeh and Contreras, 2021). Our current results lead to the hypothesis that vaccination could contribute to a part of the selection pressures on SARS-CoV-2, especially on the spike protein. It has been shown that, within populations, the frequency distribution of non-synonymous polymorphisms is negatively skewed relative to the distribution of synonymous polymorphisms under purifying selection (Hahn et al, 2002; Hughes, et al. 2005). Therefore, this hypothesis can be examined using a modified Tajima’s *D* statistics, which takes more negative values for non-synonymous (*D*_nonsyn_) than for synonymous sites (*D*_syn_) of spike gene under purifying selection (Hahn et al, 2002, Hughes, et al., 2005). The average number of pairwise synonymous differences (*k*S) and the average number of nonsynonymous pairwise differences (*k*N), the number of synonymous segregating sites (S_S_), and the number of nonsynonymous segregating sites (S_N_) were computed with equation as Austin Hughes described (1) S*_S_=S_S_/a1, (2) S*_N_=S_N_/a1, where a1 is as 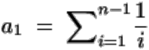 (Tajima, 1989, Hughes, et al., 2005). *D*_syn_ was defined as *k*S − S*_S_, divided by the standard error of that difference, and *D*_nonsyn_ was defined as *k*N − S*_N_, divided by the standard error of that difference (Tajima, 1989, Hughes, et al., 2005). The values of K_a_/K_s_ (the ratio of the number of nonsynonymous substitutions per non-synonymous site (K_a_), to the number of synonymous substitutions per synonymous site (K_s_), in a given period of time), and the neutrality index of the McDonald-Kreitman (MK) test of the spike protein (RaTG13 as the outgroup sequence) were analyzed by DNASP6 software (Rozas, et al., 2017).

Unlike Tajima’s *D* test, it has been demonstrated that *D*_nonsyn_ and *D*_syn_ analysis is independent of sample size, so the value of *D*_nonsyn_ or *D*_syn_ can be compared between data sets of different sizes (Hughes et al., 2008). The same excess of low-frequency alleles in non-synonymous polymorphism was also observed (*D*_nonsyn_ = -1.482 to −2.714; *P* < 0.01). In addition, the K_a_/K_s_ values were less than 1 and the neutrality index values of the MK test were significantly more than 1 (*P*<0.05) in all countries (Table 1). These results confirmed that purifying selection led to constraint on the available neutral mutations at non-synonymous sites of the spike gene of delta variants, consistent to previous reports of SARS-CoV-2 variants in 2020 (Chaw et al, 2020).

Because non-synonymous and synonymous mutations are affected unequally by purifying selection, *D*_nonsyn_ is expected to be disproportionally lower than *D*_syn_ (Hahn et al., 2002). It has been proposed that the value of Δ*D* (*D*_syn_-*D*_nonsyn_) increased as purifying selection became stronger, and Δ*D* (*D*_syn_-*D*_nonsyn_) is also able to rule out the homogenous effects (population expansion, selective sweep etc.) in neutrality tests (Hahn et al, 2002). We observed that Δ*D* of the spike gene of SARS-CoV-2 delta variants is positively proportional to fully vaccination coverage rate (*R*^2^= 0.505, *P*=0.001). When using *D* ratio (*D*_nonsyn_/*D*_syn_), the standard error terms in both equations of *D*_nonsyn_ and *D*_syn_ cancel out the sample size effect (Hughes, 2008). The positive correlation of *D* ratio and fully vaccination rate became even more evident (*R*^2^= 0.723, *P*<10^−4^) (Figure 2). Therefore, purifying selection pressure of SARS-CoV-2 spike gene increased as the vaccination coverage rate increased. This suggests that vaccination, in combination with other mitigation strategies, plays an important role in the purifying selection force of SARS-CoV-2 spike protein.

**Figure 2.**
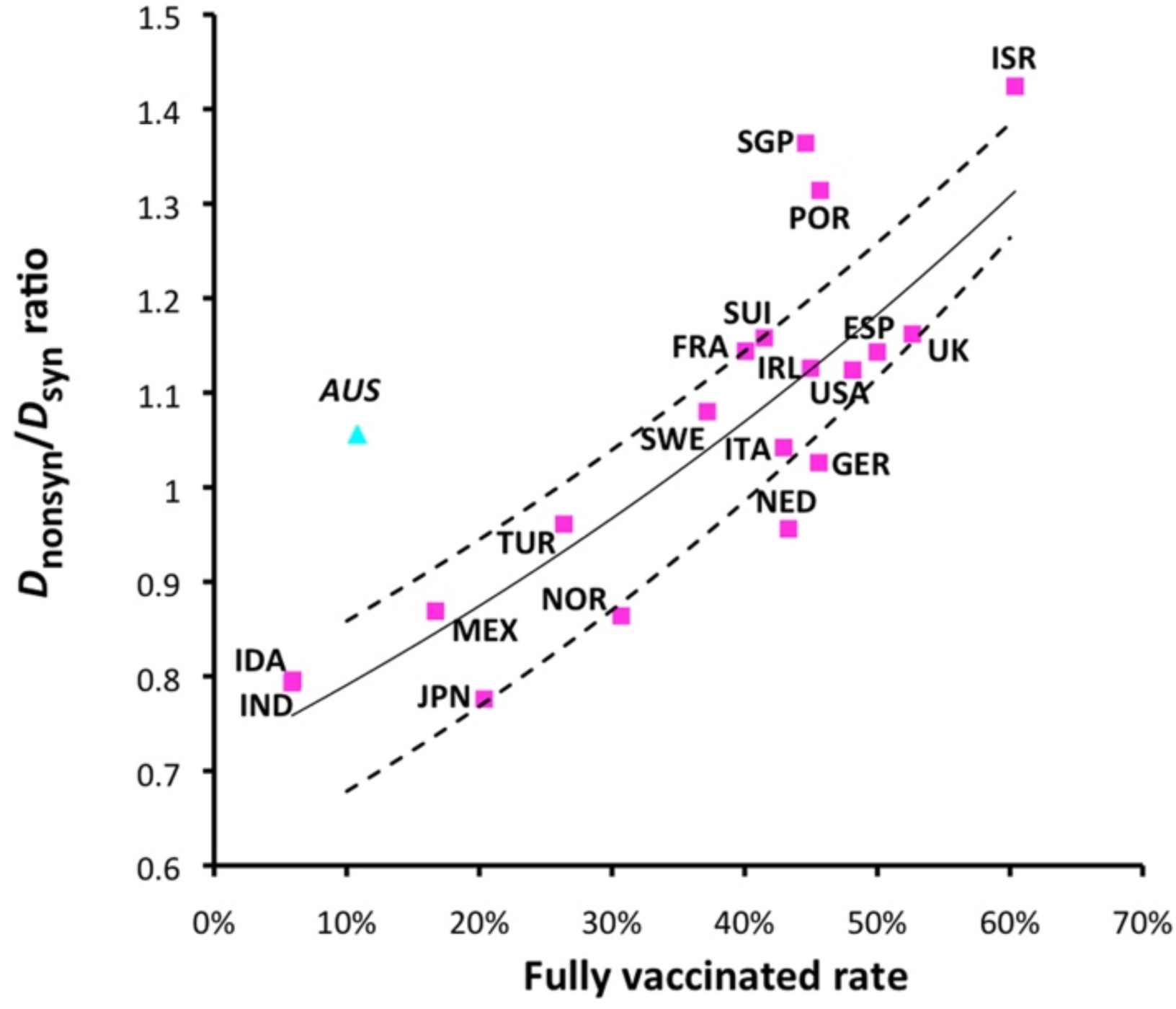
Correlation between full vaccinated rate and *D* ratio (*D*_nonsyn_ /*D*_syn_) of the spike gene of SARS-CoV-2 delta variants. Exponential regression (solid) line was draw based on 19 countries (pink square) with a calculated 95% confidence interval (dashed lines). Australia (AUS) is labeled as outliers.

### Application of neutrality tests and their limitations

Neutrality tests are very powerful tools to study population genetics and viral evolution. However, these tests require careful interpretation due to two known limitations in this study: (1) sampling across countries and the representation of sequences in public databases, (2) the timing of introductions and vaccination campaigns. Previously we have applied various neutrality tests to analyze SARS-CoV-2 transmission and evolution in Diamond Princess cruise (Yeh and Contreras, 2021). In contrast to the shipboard quarantine, where the virus spread in a closed environment, many more complicated factors (mitigation efforts, travel, herd immunity, etc.) could influence viral evolution during pandemics among different countries.

Bhatt et al. have shown that the Tajima *D* test performs very well with low type I error rate (less than 5%) when the θ value (the substitutions per site of a sequence alignment) is between 0.1 to 10^−4^ in different RNA viruses (Bhatt et al., 2010). Because θ values in this study ranged between 0.0009 to 0.032, the type I error rate is low. The Tajima *D* test is also much less influenced by RNA recombination, which we have reported previously (Yeh and Contreras, 2020, Yeh and Contreras, 2021). Tajima *D* test results depend on the interplay of many parameters: positive and negative selection, population size dynamics, spatial structure and migration, random genetic drift, etc. A key limitation of the Tajima *D* test is the difficulty to separate the effects of each parameter. However, the selection pressure and demographic expansion can be verified in combination with other neutrality tests. Our results of *DH* and Zeng’s *E* test suggests that purifying selection, not demographic expansion, was the acting force. Tajima *D* may show negative values due to a recent bottleneck event rather than selection, but the bottleneck effect should impact all types of polymorphism equally (Tajima, 1989). Significant differences between *D*_nonsyn_ and *D*_syn_ (either Δ*D* or *D* ratio (*D*_nonsyn_/*D*_syn_)), which efficiently eliminated the homogenous effects, strongly indicates that purifying selection was at play and is positively corelated with vaccination coverage rate within these samples.

The inter-species divergence K_a_/K_s_ test has been shown to be poorly equipped to detect positive selection in short time scales for the comparison of young evolutionary lineages like SARS-CoV-2 (Mugal, et al., 2014). Consistent with other analytical attempts, our K_a_/K_s_ data was inconclusive (Table 1). This has been the case with other applications of the K_a_/K_s_ test in SARS-CoV-2 spike gene, which resulted in extreme values and controversial results (Kang et al., 2021), In contrast to K_a_/K_s_, the MK test cannot be used to locate specific sites under selection, although this test enables us to estimate the adaptive substitution rate of a gene (Smith and Eyre-Walker, 2002). Variants of the MK test have been used to analyze the evolution of RNA viruses (Bhatt et al., 2010). Since the MK test relies on the comparison of the ratio of polymorphism to fixed differences of both synonymous and nonsynonymous mutations, selection of an outgroup sequence is critical for this test. The number of fixed synonymous mutations may be underestimated in a rapidly evolving virus like SARS-CoV-2 when a distant outgroup is applied (Baudry and Depaulis, 2003). Since the K_a_/K_s_ or the MK tests may not be the appropriate tests for SARS-CoV-2 delta variants, we only included them to confirm the observation of purifying selection of SARS-CoV-2 spike gene here as well. We also did not include neutrality tests based on haplotype distribution or linkage disequilibrium, because they are expected to be strongly affected by RNA recombination that commonly occurred in SARS-CoV-2 populations (Yeh and Contreras, 2020, Ignatieva et al, 2021, Yeh and Contreras, 2021).

### Perspectives

More virulent strains of SARS-CoV-2 have emerged with enhanced transmissibility and immune evasion properties. For example, multiple variants have escaped neutralizing antibodies developed to target the spike protein receptor-binding or N-terminal domain (Harvey, et al., 2021). What’s more, the case numbers of breakthrough infection caused by the delta variant have increased drastically worldwide (Farinholt et al., 2021).

Of all countries studied, our observations show that higher vaccination rates corresponded with lower mutation frequencies and higher *D*_nonsyn_/*D*_syn_ values for the whole genome and the spike gene. We conclude then, that as the vaccination coverage rate increases, purifying selection forces of nonsynonymous mutations also increase.

Thus, we recommend that: 1) universal vaccination should be administered as soon as possible to suppress the generation of deadly mutations; 2) mitigations strategies such as personal protection equipment, social distancing, etc., should be continued until full vaccination is reached to prevent viral transmission; 3) genomic surveillance should be undertaken to monitor for new mutations; and 4) more sequence summary statistics of RNA virus evolution are required to facilitate understanding new COVID-19 outbreaks.

## Data Availability

1.The World Health organization. Tracking SARS-CoV-2 variants.
2. Global Initiative on Sharing All Influenza Data.
3. Coronavirus (COVID-19) Vaccinations. Our World in Data.

https://www.who.int/en/activities/tracking-SARS-CoV-2-variants/

https://www.gisaid.org/

https://ourworldindata.org/covid-vaccinations

## Author contributions

All authors contributed to study concept, rationale, and initial manuscript drafts, interpretation of data, and final manuscript preparation. All authors have read and approved the final version of the manuscript.

## Funding

No funding

### Ethical approval

None declared.

## References

Bhatt, B., Katzourakis, A., Pybus, O.G., (2010) Detecting natural selection in RNA virus populations using sequence summary statistics. Infect Genet Evol., 10, 421–30.

Baudry, E., Depaulis, F. (2003) Effect of misoriented sites on neutrality tests with outgroup. Genetics, 165, 1619–1622

Chaw, S.M., Tai, J.H., Chen, S.L., Hsieh, C.H., Chang, S.Y., Yeh, S.H., Yang, W.S., Chen, P.J., Wang, H.Y. (2020) The origin and underlying driving forces of the SARS-CoV-2 outbreak. J Biomed Sci., 27, 73.

Farinholt, T., Doddapaneni, H., Qin, X., Menon, V., Meng, Q., Metcalf, G., Chao, H., Gingras M., Farinholt, P., Agrawal, C., Muzny, D.M., Piedra, P.A., Gibbs, R.A., Petrosino, J. (2021) Transmission event of SARS-CoV-2 Delta variant reveals multiple vaccine breakthrough infections. medRxiv doi: 10.1101/2021.06.28.21258780.

Harvey, W.T., Carabelli, A.M., Jackson, B., Gupta, R.K., Thomson, E.C., Harrison, E.M., Ludden, C., Reeve, R., Rambaut, A., COVID-19 Genomics UK (COG-UK) Consortium, Peacock S.H., Robertson D.L. (2021) SARS-CoV-2 variants, spike mutations and immune escape. Nat Rev Microbiol.,19, 409–424.

Hahn, M.W., Rausher, M.D., Cunningham, C.W. (2002) Distinguishing between selection and population expansion in an experimental lineage of bacteriophage T7. Genetics, 161, 11–20.

Hughes, A.L., (2005) Evidence for Abundant Slightly Deleterious Polymorphisms in Bacterial Populations. Genetics, 169, 533–538.

Hughes, A.L., Friedman, R., Rivailler, P., French, J.O., (2008) Synonymous and nonsynonymous polymorphisms versus divergences in bacterial genomes. Mol Biol Evol.,25, 2199–2209.

Ignatieva, A., Hein, J., Jenkins, P.A, (2021) Evidence of ongoing recombination in SARS-CoV-2 through genealogical reconstruction. bioRxiv, https://doi.org/10.1101/2021.01.21.427579

Kang, L., He, G., Sharp, A.K., Wang, X., Brown, A.M., Michalak. P., Weger-Lucarelli, J. (2021) A selective sweep in the Spike gene has driven SARS-CoV-2 human adaptation. Cell, 184, 4392–4400.

Kirzinger, A., Sparks, G., Brodie, M. (2021) KFF COVID-19 vaccine monitor-Rural America. https://www.kff.org/coronavirus-covid-19/poll-finding/kff-covid-19-vaccine-monitor-in-their-own-words-six-months-later/ Accessed 13 July 2021.

Mugal, C.F., Wolf, J.B., Kaj, I. (2013) Why time matters: codon evolution and the temporal dynamics of dN/dS. Mol Biol Evol., 31, 212–231.

Rozas, J., Ferrer-Mata, A., Sánchez-DelBarrio, J.C., Guirao-Rico, S., Librado, P., Ramos-Onsins, S.E., Sánchez-Gracia, A. (2017) DnaSP 6: DNA Sequence Polymorphism Analysis of Large Data Sets. Mol Biol Evol., 34, 3299–3302.

Roy, C., Mandal, S.M., Mondal, S.K., Mukherjee, S., Mapder, T., Ghosh, W., Chakraborty, R. (2020) Trends of mutation accumulation across global SARS-CoV-2 genomes: Implications for the evolution of the novel coronavirus. Genomics, 112, 5331–5342.

Schmelz, K., Bowles, S., (2021) Overcoming COVID-19 vaccination resistance when alternative policies affect the dynamics of conformism, social norms, and crowding out. Proc Natl Acad Sci U S A, 118, e2104912118.

Smith, N.G., Eyre-Walker, A., (2002) Adaptive protein evolution in Drosophila. Nature, 415, 1022–1024.

Tajima, F. (1989) Statistical method for testing the neutral mutation hypothesis by DNA polymorphism. Genetics, 123, 585–595.

Yeh, T.Y., Contreras, G.P. (2020) Emerging viral mutants in Australia suggest RNA recombination event in the SARS-CoV-2 genome. Med J Aust, 213, 44–44.e1.

Yeh, T.Y., Contreras, G.P. (2021) Viral transmission and evolution dynamics of SARS-CoV-2 in shipboard quarantine. Bull World Health Organ., 99, 486–495.

Zeng, K., Fu, Y.X., Shi, S., Wu, C.I. (2006) Statistical tests for detecting positive selection by utilizing high-frequency variants. Genetics, 174, 1431–1439.

